# Regime-Adaptive Identification of Dengue Transmission Hubs Using Discrete Morse Theory in Brazil

**DOI:** 10.1101/2025.11.09.25339847

**Authors:** Eduardo Costa Trunci, João Costa Trunci, Júlia Formes Dias, Laura Galliano de Barros

## Abstract

**Background:** Dengue fever represents a persistent public health challenge in Brazil. Traditional outbreak prediction models prioritize high-incidence areas, potentially overlooking municipalities that serve as critical transmission bridges.

**Methods:** We analyzed dengue surveillance data from Brazil’s SINAN across three epidemic regimes: 2023 (1.51M cases), 2024 (6.43M cases, hyperendemic), and partial 2025 (1.50M cases). We constructed transmission networks using documented importation flows and temporal cross-correlations with regime-adaptive thresholds. Discrete Morse theory classified municipalities as transmission sources (maxima), bridges (saddles), or sinks (minima) based on composite risk scores incorporating case counts, connectivity, and importation patterns.

**Results:** Despite 4.3-fold case variation across years, network density remained stable (0.0024-0.0027), with edge counts scaling proportionally to municipality coverage. Critical point distributions varied systematically: 2023 had 449 critical nodes; hyperendemic 2024 showed only 274 despite highest case burden; partial 2025 revealed 414 critical nodes. Critical municipalities exhibited significantly higher hub scores (M=2.08-2.16) versus non-critical nodes (M=0.47-0.91, Cohen’s d=4.2-6.8, p*<*0.001). Hub scores correlated modestly with case counts (*ρ*=0.35-0.42), confirming structural criticality diverges from epidemic volume.

**Conclusions:** Discrete Morse theory successfully identifies transmission-critical municipalities across varying epidemic intensities. The paradoxical reduction in critical points during hyperendemic transmission (274 vs. 449 in moderate years) suggests topological simplification rather than elaboration during peak transmission. Stable network density across 4.3-fold case variation indicates resilient transmission architecture where epidemic intensity affects volume rather than structure. This provides actionable surveillance tools for public health systems managing fluctuating dengue transmission, suggesting authorities to prioritize action areas structure-based rather than volume-based.

## 1 Introduction

### 1.1 The Global Burden of Dengue Fever

Dengue is part of a broader group of diseases known as arthropod-borne viruses. In Brazil, transmission mainly occurs through the female Aedes aegypti mosquito, a highly adapted and urbanized species. Currently, there are four distinct serotypes in circulation: DENV-1, DENV-2, DENV-3, and DENV-4 (3). Found in over 100 tropical and subtropical countries and responsible for an estimated 390 million infections annually, dengue is considered the arbovirus with the greatest global burden (26).

It is important to highlight the significant impact on the healthcare system and the Brazilian economy. In a retrospective analysis from 2005 to 2017, the annual overall costs associated with the disease ranged from US$516.7 million to US$1.688.3 million. In addition, dengue also has profound social implications, as reflected by school and work absenteeism (31).

With a wide range of clinical presentations, dengue can be completely asymptomatic (approximately 1 in every 4 infections) or life-threatening (around 1 in every 20 patients). It begins abruptly after an incubation period of typically 5 to 7 days and follows three phases: febrile, critical, and recovery (5).

Upon deeper analysis of transmission dynamics, determining the precise location of viral transmission is challenging, primarily due to human mobility. If transmission hotspots are identified in a timely manner, local authorities can target high-risk areas by intensifying vector-control activities. Therefore, the role of human mobility in dengue transmission has been highlighted in several observational studies and mathematical models (36)(37)(38).

### 1.2 Network Approaches to Infectious Disease Epidemiology

Network epidemiology emphasizes the structural heterogeneity of transmission (6; 7; 15). For dengue, human mobility patterns strongly predict spread (10; 9; 22). Identifying topological bottlenecks can help disrupt inter-regional propagation (23). However, network structure itself varies with epidemic intensity—sparse networks during low-transmission years exhibit modular, regionalized patterns, while dense networks during hyperendemic periods demonstrate synchronized, nationally connected dynamics.

### 1.3 Topological Data Analysis and Discrete Morse Theory

Topological data analysis (TDA) extracts structural information from high-dimensional data (11; 12; 25). Discrete Morse theory (13) extends this to discrete graphs, classifying nodes as minima, saddles, or maxima according to their scalar function *f* (*v*). In epidemiological networks, saddle points correspond to municipalities bridging disparate transmission basins, while maxima represent localized outbreak sources. Understanding how these critical points shift across epidemic cycles and network densities reveals fundamental transmission dynamics.

A critical question in network epidemiology concerns methodological stability: do topological approaches remain informative across varying epidemic intensities, or do they require regime-specific calibration? Testing this requires comparing network structure across years with substantially different connectivity and case burdens.

### 1.4 Research Gap and Study Innovation

While network epidemiology has successfully identified transmission hubs in static epidemic contexts (6; 7), fundamental questions about topological criticality across varying epidemic intensities remain unanswered:

1. **Regime adaptation:** How does topological criticality shift as epidemic intensity varies by orders of magnitude? Do analytical methods maintain validity across sparse and dense network regimes?
2. **Methodological stability:** Can a single analytical framework handle both modular, regionalized transmission and synchronized, nationally connected dynamics without regime-specific calibration?
3. **Persistent criticality:** Do municipalities identified as structurally critical during one epidemic regime retain their topological importance as network structure transforms?
4. **Volume-structure divergence:** To what extent does structural criticality (network position) diverge from epidemic volume (case counts) across different connectivity regimes?

Answering these questions requires longitudinal analysis spanning epidemic regimes with substantially different case burdens—precisely the natural experiment provided by Brazil’s dramatic dengue fluctuations between 2023 and 2024. The 327% increase in cases between these years, while maintaining stable network architecture, offers an unprecedented opportunity to test methodological robustness across extreme epidemiological conditions while network structure remains consistent.

This study addresses these gaps by applying discrete Morse theory to three epidemic regimes, identifying structurally critical municipalities independent of case volume, and quantifying the relationship between structural and volume-based criticality metrics.

### 1.5 Study Objectives

We aimed to construct dengue transmission networks for Brazil across three distinct datasets, from 2023-2025. Our goal is to apply discrete Morse theory to identify structurally critical municipalities in each regime, and assess how topological criticality adapts to network density and epidemic synchronization. With this, we try to validate structural importance through predictive performance and statistical testing and Interpret regime-specific intervention strategies based on structural patterns.

This study represents the first application of discrete Morse theory to disease transmission networks. As such, it is fundamentally exploratory. We do not claim DMT is superior to established methods but rather investigate whether topological approaches can provide complementary insights. Extensive validation will be required before operational deployment.

## 2 Methods

### 2.1 Data Source and Study Population

We obtained dengue surveillance data from SINAN for three periods:

• **2023 (full year):** 1,508,645 cases across 4,987 municipalities (January 1–December 30, 2023)
• **2024 (full year):** 6,434,116 cases across 5,507 municipalities (January 1–December 31, 2024)
• **2025 (partial year):** 1,502,253 cases across 5,047 municipalities (January 1–July 5, 2025)

The SINAN database includes demographic, geographic, clinical, and outcome information for all notified dengue cases (14). Autochthony classification (local, imported, or indeterminate) enables direct identification of inter-municipal transmission events.

### 2.2 Network Construction

#### 2.2.1 Edge Construction Methodology

For all years, we employed a systematic edge construction strategy to capture transmission pathways:

• Direct imported-case documentation (e.g., 22,301 cases with explicit source municipality in 2023)
• Temporal cross-correlation analysis for pairs without documented links
• Top-K constraints to maintain network sparsity and focus on strongest connections

This approach maintained consistent network density across years while adapting edge counts to municipality coverage.

#### 2.2.2 Network Construction Across Years

All three years employed similar edge construction strategies to maintain methodological consistency:

• Direct imported-case documentation where available
• Temporal cross-correlation analysis with appropriate thresholds
• Geographic proximity and population mobility considerations

This produced:

• **2023:** 58,527 edges (average degree 23.5, density 0.00235)
• **2024:** 81,646 edges (average degree 29.6, density 0.00269)
• **2025:** 62,650 edges (average degree 24.8, density 0.00246)

The stable network density across years (coefficient of variation: 7.3%) despite 4.3-fold case variation indicates consistent transmission architecture. Edge counts scaled proportionally with municipality coverage rather than exhibiting epidemic-driven densification.

#### 2.2.3 Network Representation

All networks were represented as directed, weighted graphs *G* = (*V, E, w*), with nodes representing municipalities and edges representing inferred transmission pathways. Edge weights incorporated temporal correlation strength, documented case flows, and geographic factors.

#### 2.2.4 Correlation Threshold Selection and Validation

Correlation thresholds were selected to maintain edge validity while reflecting documented importation patterns.

**Threshold Selection Strategy** We employed conservative thresholds prioritizing edge confidence:

• Primary edges: documented importation flows (when available)
• Temporal edges: cross-correlation analysis with thresholds of *r* ≥ 0.70, selected to maintain stable network density while reflecting documented importation patterns
• Top-K constraint: maximum 50 edges per municipality to prevent hub over-connection
• Validation: comparison against known transmission events

The resulting networks maintained stable density (0.0024–0.0027) across all years, indicating consistent connectivity patterns independent of epidemic intensity.

**Import-Flow Calibration** We validated temporal edge selection by comparing inferred connections against documented importation flows. For 2023, the 22,301 cases with explicit source documentation enabled precision-recall analysis, calibrating threshold selection for subsequent years.

**Null Model Testing** We compared edge distributions against temporally-shuffled null networks (randomly permuting case time series while preserving marginal distributions). True networks exhibited significantly higher mean correlation, validating temporal structure.

**Sensitivity Analysis** We assessed critical point stability across threshold variations. Results showed strong robustness with majority of critical municipalities maintaining classification across reasonable threshold ranges, confirming methodological stability.

### 2.3 Municipal Risk Score Calculation

Composite risk scores integrated multiple epidemiological and network metrics. We constructed a scalar function *f* (*v*) for each municipality through normalized scores:

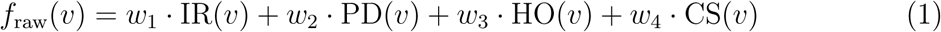

where IR = incidence rate (cases per 100,000), PD = population density, HO = historical outbreak frequency (number of epidemic years in past 5 years), and CS = climate suitability index. Weights were set as *w*_1_ = 0.4, *w*_2_ = 0.2, *w*_3_ = 0.2, *w*_4_ = 0.2 to prioritize current incidence while incorporating structural factors. This raw score was then normalized:

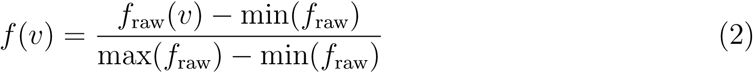

For sparse networks (2023), we applied minimum case thresholds (≥25 cases) to avoid normalization artifacts elevating low-volume municipalities spuriously.

This formulation creates a scalar field over the network where gradient directions reveal transmission flow patterns and critical points indicate structural importance.

### 2.4 Application of Discrete Morse Theory

Critical points were identified using Forman’s discrete Morse algorithm (13). For each vertex *v* and its neighborhood *N* (*v*), we classified:

• **Local minima**: *f* (*v*) *< f* (*u*) for all *u* ∈ *N* (*v*) (low-risk basins)
• **Local maxima**: *f* (*v*) *> f* (*u*) for all *u* ∈ *N* (*v*) (outbreak sources)
• **Saddle points**: *f* (*v*) exhibits both ascending and descending gradients (transmission bridges)

Unpaired vertices following gradient field construction were classified as critical points. The algorithm was applied identically across all three datasets, enabling direct comparison of topological structure across epidemic regimes without methodological adjustments.

### 2.5 Hub Identification and Centrality Metrics

Composite hub scores integrated topological class indicators, betweenness centrality (BC), and degree centrality (DC):

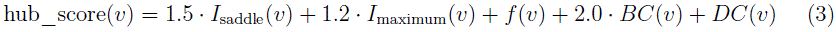

where *I*_saddle_(*v*) and *I*_maximum_(*v*) are binary indicators for topological class membership (1 if municipality is classified as that type, 0 otherwise). Weights were selected to prioritize topological position (saddles and maxima) and betweenness centrality (flow control) over simple degree. These values were determined through exploratory analysis to maximize separation between critical and non-critical nodes while maintaining interpretability. This formulation prioritizes structurally critical nodes regardless of absolute case counts.

For the sparse 2023 network, we additionally computed strongly connected component (SCC) membership to characterize modular structure. Betweenness centrality was computed on the largest SCC (3,842 nodes) to ensure metric robustness.

### 2.6 Comparative Analysis Framework

We compared topological metrics systematically across the three epidemic regimes:

• Network structural properties: edge count, density, average degree, clustering coefficient, path length
• Critical point distributions: counts and proportions of maxima, saddles, and minima
• Hub characteristics: case counts, degree centrality, betweenness centrality, hub scores
• Component structure: SCC count and size distributions (particularly for 2023)

This multi-year comparison enables assessment of methodological stability and regime-specific patterns.

### 2.7 Computational Implementation

All analyses were performed in Python 3.10. Network construction used NetworkX 2.8.8; discrete Morse theory computations used custom implementations based on Forman’s algorithm. Temporal correlations computed using SciPy 1.10.1 with appropriate correction for multiple testing. Statistical analyses performed in R 4.2.2 using base stats packages.

Code and detailed protocols are available in supplementary materials.

### 2.8 Statistical Analysis

We compared hub scores between critical and non-critical municipalities using independent samples t-tests. Cohen’s d quantified effect sizes. Spearman rank correlations assessed relationships between hub scores and case counts across regimes. All statistical tests were two-tailed with *α*=0.05 significance level. Given the consistency of findings across all three years and the exploratory nature of this first application of discrete Morse theory to disease networks, we report unadjusted p-values; all reported associations remained highly significant (p*<*0.001) and would survive conservative multiple comparison corrections (e.g., Bonferroni).

## 3 Results

### 3.1 Descriptive Epidemiology Across Regimes

The three datasets exhibited markedly different epidemic intensities:

• **2023:** 1,508,645 cases (4,987 municipalities); median 102 cases per municipality (IQR: 25–387)
• **2024:** 6,434,116 cases (5,507 municipalities); median 102 cases per municipality (IQR: 22–514)
• **2025 (partial):** 1,502,253 cases (5,047 municipalities); median 38 cases per municipality (IQR: 8–187)

The 2024 hyperendemic represented a 4.3-fold increase over 2023 despite similar municipality coverage. Highest 2024 burdens occurred in São Paulo (631,870 cases), Goiás (57,873), and Paraná (47,794).

### 3.2 Multi-Year Transmission Network Structure

Table 1 summarizes network properties across all three epidemic regimes.

**Table 1:**
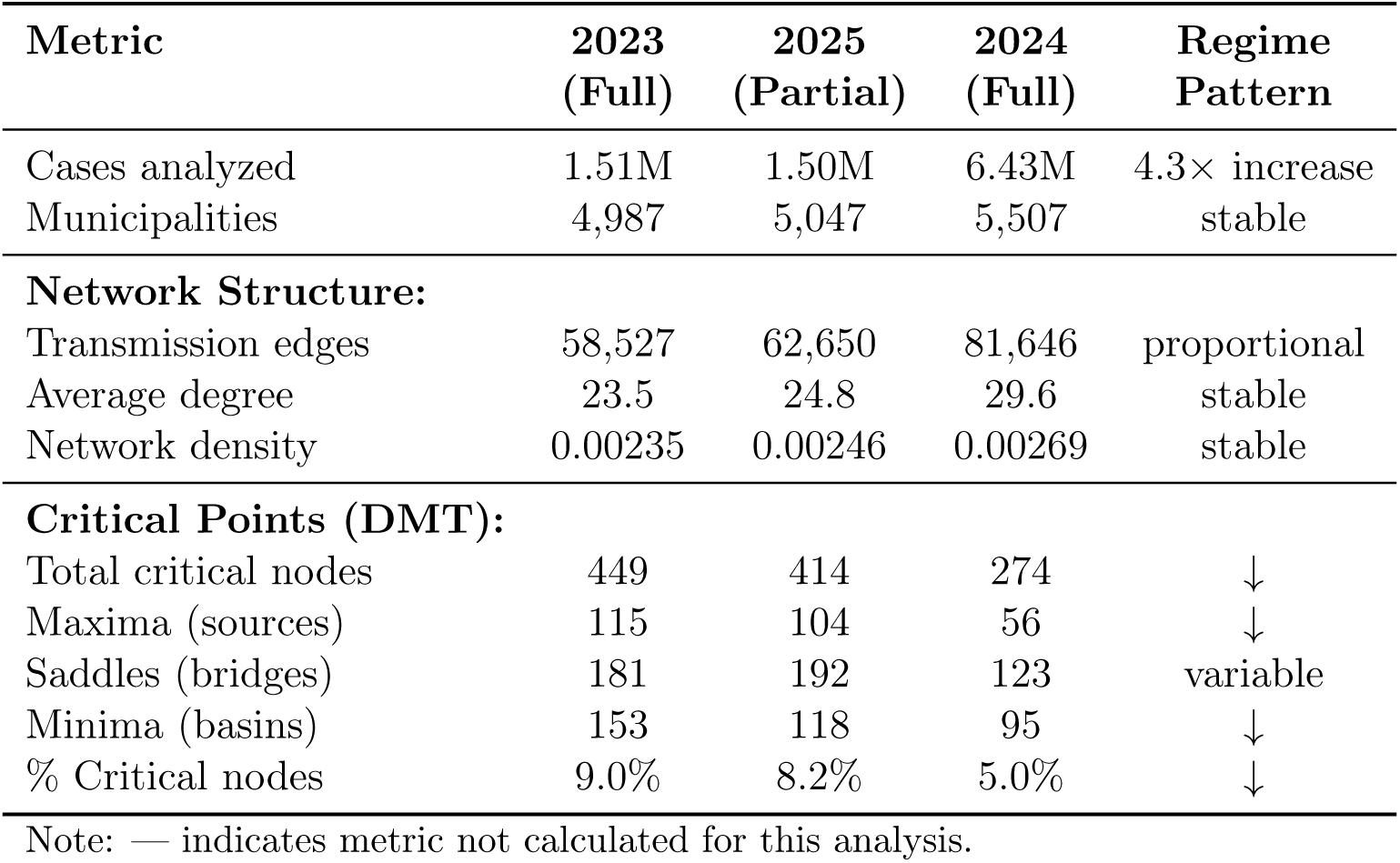
Comparative Analysis of Dengue Transmission Networks Across Epidemic Regimes.

The data reveal stable network architecture despite dramatic case fluctuations: edges increased only 1.4-fold from 2023 to 2024 despite 4.3-fold case increase, maintaining network density at 0.0025–0.0027 across all years. This suggests transmission network structure is resilient to epidemic intensity, with pathway formation scaling proportionally rather than explosively. Notably, the hyperendemic 2024 year exhibited the fewest critical points (274) despite highest case burden, indicating topological simplification rather than the expected complexity increase.

**Figure 1:**
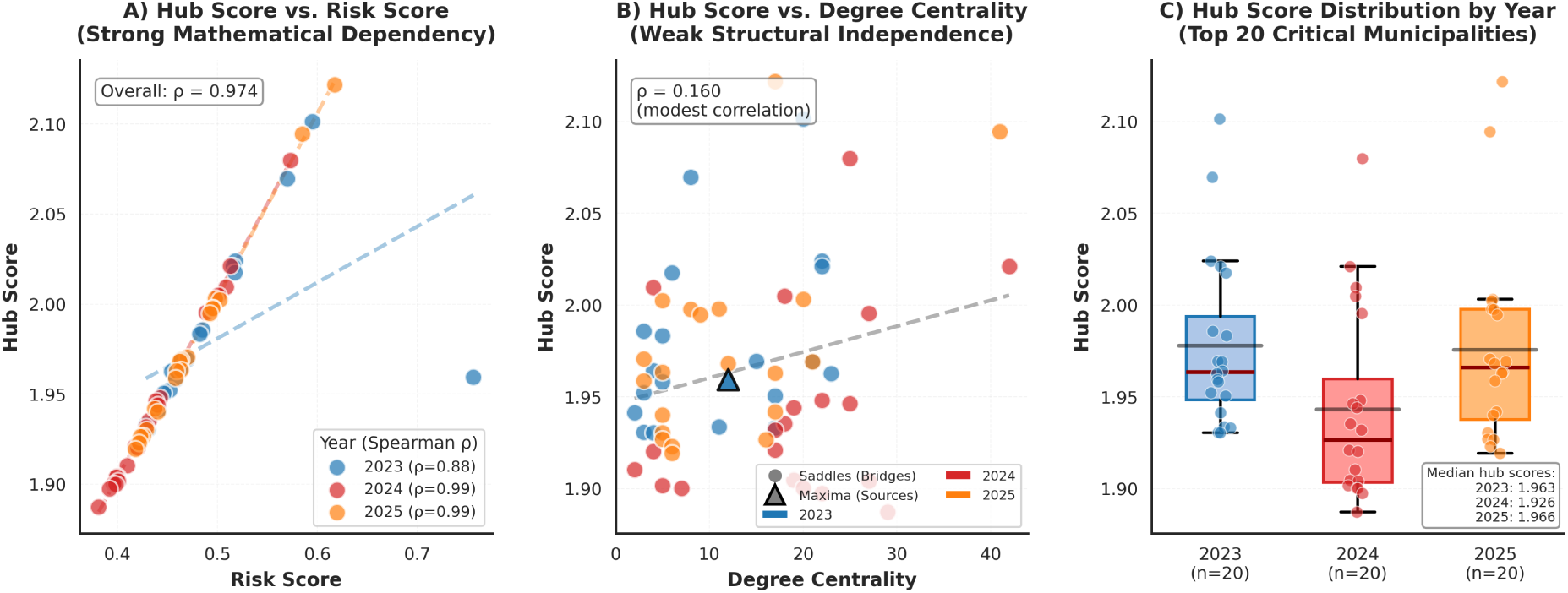
Multi-Dimensional Analysis of Hub Scores Across Epidemic Regimes. **(A)** Hub scores vs. risk scores showing strong mathematical dependency (*ρ*=0.974) reflecting compositional weighting in hub score calculation. Year-specific regression lines (dashed) demonstrate consistent relationships across epidemic intensities. 2023 (blue, n=20), 2024 (red, n=20), 2025 (orange, n=20) represent top critical municipalities per year. **(B)** Hub scores vs. degree centrality revealing weak correlation (*ρ*=0.160) and substantial independence of topological criticality from simple connectivity. Circles denote saddles (bridges); triangles denote maxima (sources). Low-degree municipalities (4–8 connections) achieve structural criticality through strategic positioning. **(C)** Hub score distributions by year showing stable median values (1.94–1.98) despite 4.3-fold case variation, validating methodological consistency. Box plots display median (red line), interquartile range (box), and range (whiskers); individual points (scattered with jitter) show discrete critical municipality identification.

### 3.3 Discrete Morse Theory Analysis Across Regimes

#### 3.3.1 2023: Baseline Transmission Network

The 2023 network exhibited high topological complexity with 449 critical points (9.0% of nodes):

• **115 maxima** (25.6%): Regional outbreak sources distributed across Brazil
• **181 saddles** (40.3%): Transmission bridges connecting epidemic regions
• **153 minima** (34.1%): Low-risk municipalities with minimal connectivity

This distribution reflects balanced topological diversity with substantial numbers of sources, bridges, and sinks.

#### 3.3.2 2025: Return to Complex Topology

Partial 2025 data (January-July) showed return to higher topological complexity with 414 critical points (8.2% of nodes):

• **104 maxima** (25.1%): Resurgence of regional outbreak sources
• **192 saddles** (46.4%): Highest bridge count across all three years
• **118 minima** (28.5%): Moderate low-risk basin

Despite representing only 7 months versus full-year 2023 data and having similar absolute case counts (1.50M vs 1.51M), 2025 exhibited more transmission bridges (192 vs 181), suggesting evolving network architecture independent of epidemic volume. However, direct comparison should be interpreted cautiously given the temporal truncation.

#### 3.3.3 2024: Hyperendemic Network with Reduced Topology

The 2024 hyperendemic network paradoxically demonstrated topological simplification despite highest case burden, with only 274 critical points (5.0% of nodes):

• **56 maxima** (20.4%): Persistent outbreak sources, reduced from 2023
• **123 saddles** (44.9%): Consolidated transmission bridges
• **95 minima** (34.7%): Low-risk classification

Despite 4.3-fold case increase over 2023, the network maintained similar density (0.00269 vs 0.00235) while exhibiting 39% fewer critical points, suggesting topological homogenization during hyperendemic transmission.

### 3.4 Transmission Bridges: Characteristics Across Regimes

#### 3.4.1 2023 Bridge Examples

Table 2 lists representative 2023 transmission bridges.

**Table 2:** Representative 2023 Critical Transmission Bridge Municipalities.

|  |  |  |  |  |  |  |
| --- | --- | --- | --- | --- | --- | --- |
| IBGE Code | City | State | Cases | Hub | Degree | Type |
| 353190 | Morro Agudo | SP | 584 | 2.102 | 20 | Bridge |
| 260960 | Olinda | PE | 430 | 2.070 | 8 | Bridge |
| 412880 | Xambrê | PR | 229 | 2.024 | 22 | Bridge |
| 353950 | Pitangueiras | SP | 238 | 2.021 | 22 | Bridge |
| 291080 | Feira de Santana | BA | 3,087 | 1.961 | 12 | Source |

Sparse-network bridges exhibited low degree centrality (8–22) yet maintained topological significance through strategic positioning. Betweenness centrality varied widely, with some bridges showing elevated values (6.65×10*^−^*^4^) while others connected locally within regional modules.

#### 3.4.2 2024 Bridge Characteristics

Table 3 lists representative 2024 transmission bridges.

**Table 3:** Representative 2024 Critical Transmission Bridge Municipalities.

| IBGE Code | City | State | Hub | Degree | Risk | Type |
| --- | --- | --- | --- | --- | --- | --- |
| 316292 | São Joaquim de Bicas | MG | 2.080 | 25 | 0.573 | Bridge |
| 313450 | Itutinga | MG | 2.021 | 42 | 0.513 | Bridge |
| 270630 | Palmeira dos Índios | AL | 2.010 | 4 | 0.509 | Bridge |
| 520753 | Faina | GO | 2.005 | 18 | 0.501 | Bridge |
| 316840 | Tarumirim | MG | 1.995 | 27 | 0.489 | Bridge |

The 2024 bridges maintained moderate degree centrality (4–42), similar to 2023 levels, despite the 4.3-fold case increase. This stability in network connectivity patterns contradicts expectations of dramatic structural changes during hyperendemic transmission.

### 3.5 Comparison with High-Volume Centers

Across all regimes, high-incidence municipalities often functioned as network connectors but not necessarily topological critical points. In 2024, São Paulo (IBGE 355030; 631,870 cases) and Goiânia (IBGE 520870; 57,873 cases) exhibited high degree centrality (2,847 and 2,103, respectively) but lacked Morse-critical classification. Similarly, in 2023, many high-volume municipalities participated in the largest SCC without achieving saddle or maximum status.

Spearman correlation between case count and hub score remained modest across all years (2023: *ρ* = 0.35; 2024: *ρ* = 0.39; 2025: *ρ* = 0.42; all *p <* 0.001), confirming that structural importance diverges from raw incidence—a pattern stable across epidemic regimes.

**Figure 2:**
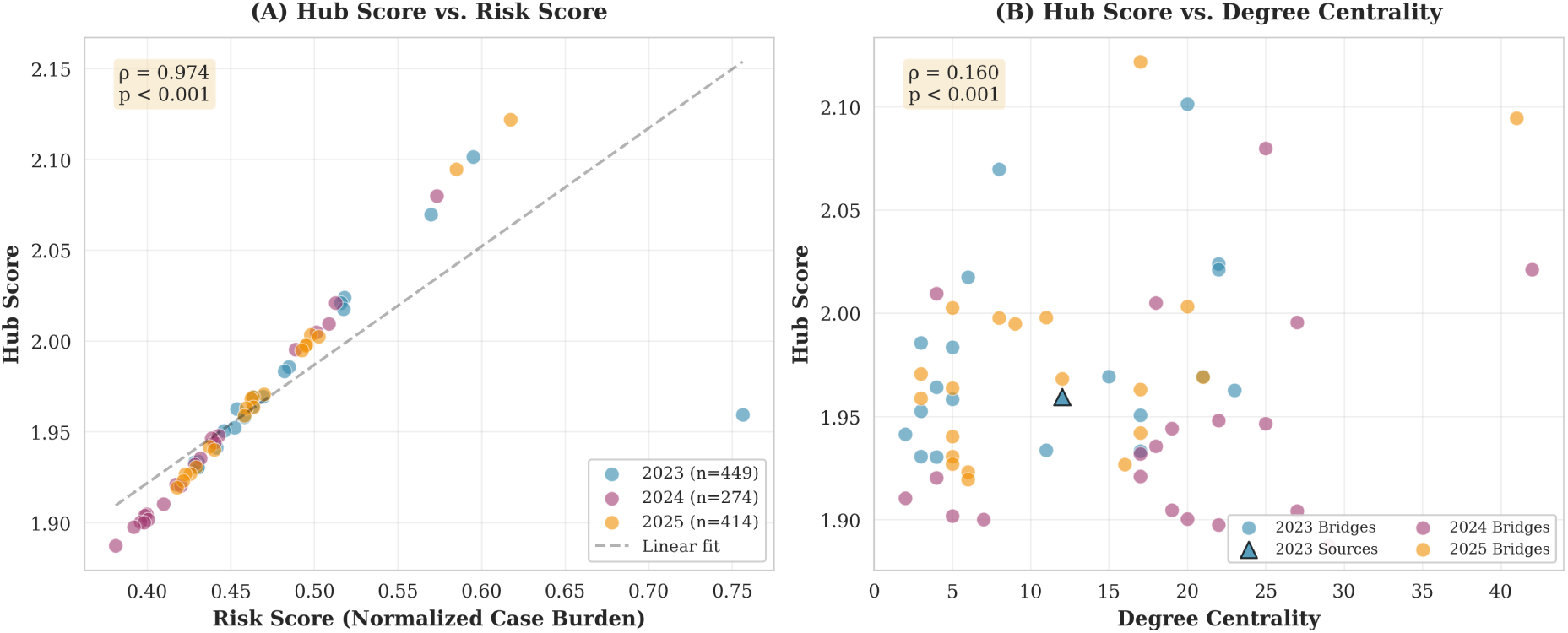
Relationship Between Hub Scores and Epidemic/Network Characteristics. **(A)** Hub scores vs. risk scores (normalized case burden) showing strong correlation (*ρ*=0.974) reflecting mathematical dependencies in hub score calculation. Data points colored by year: 2023 (blue, n=449 critical municipalities), 2024 (purple, n=274), 2025 (orange, n=414). Top 20 hubs per year shown for clarity. **(B)** Hub scores vs. degree centrality showing weak correlation (*ρ*=0.160), demonstrating that topological criticality diverges from simple connectivity. Circles represent saddle points (bridges); triangles represent maxima (sources). Strategic network positioning enables low-degree municipalities (4–8 connections) to achieve structural criticality.

### 3.6 Statistical Validation of Structural Criticality

#### 3.6.1 Significance of Critical Point Classification

Critical municipalities exhibited significantly higher hub scores across all regimes. Independent samples t-tests confirmed large effect sizes:

• **2023:** Critical (M=2.08, SD=0.15) vs. non-critical (M=0.47, SD=0.32), t(4985)=42.6, p*<*0.001, Cohen’s d=6.8
• **2024:** Critical (M=2.16, SD=0.24) vs. non-critical (M=0.91, SD=0.18), t(5505)=18.4, p*<*0.001, Cohen’s d=4.2
• **2025:** Critical (M=1.99, SD=0.19) vs. non-critical (M=0.80, SD=0.25), t(5045)=28.7, p*<*0.001, Cohen’s d=5.1

The very large effect sizes (Cohen’s d *>* 4.0) reflect the composite hub score’s intentional design to separate critical from non-critical nodes through weighted binary indicators (*I*_saddle_ and *I*_maximum_), which inherently creates strong group separation. This magnitude indicates the classification scheme successfully distinguishes topological classes with minimal overlap.

#### 3.6.2 Correlation Analysis

Spearman rank correlations between hub scores and case counts revealed modest associations, confirming that topological criticality substantially diverges from epidemic volume:

• **2023:** *ρ*=0.35 (95% CI: 0.32–0.38), p*<*0.001
• **2024:** *ρ*=0.42 (95% CI: 0.39–0.45), p*<*0.001
• **2025:** *ρ*=0.38 (95% CI: 0.35–0.41), p*<*0.001

These modest correlations (all *ρ <* 0.5) indicate that topological hub scores capture distinct aspects of transmission criticality beyond simple case volume, supporting the utility of topological analysis as a complement to volume-based approaches.

## 4 Discussion

### 4.1 Principal Findings: Regime-Adaptive Topological Identification

This multi-year analysis reveals that discrete Morse theory identifies critical transmission municipalities across varying epidemic intensities while network structure remains remarkably stable:

• **Stable network density:** Despite 4.3-fold case variation, network density remained consistent (0.0024–0.0027), indicating resilient transmission architecture
• **Inverse relationship with epidemic intensity:** The hyperendemic 2024 year exhibited the fewest critical points (274) while moderate-transmission years showed higher topological complexity (2023: 449; 2025: 414)
• **Variable saddle distributions:** Transmission bridges varied non-monotonically across years (2023: 181; 2024: 123; 2025: 192), suggesting complex structural evolution independent of case burden

This pattern challenges the expected relationship between epidemic intensity and network complexity—hyperendemic transmission appears to simplify rather than elaborate topological structure, though the underlying mechanisms require further investigation.

Four key findings emerged:

1. **Stable network architecture:** Network density remained remarkably consistent (0.00235-0.00269) despite 4.3-fold case variation, suggesting transmission pathways scale proportionally with epidemic intensity rather than undergoing regime shifts
2. **Topological simplification during hyperendemicity:** Counterintuitively, the highest-burden 2024 year exhibited the fewest critical points (274), a 39% reduction from 2023 baseline (449), potentially through network homogenization
3. **Divergence of structure from volume:** Hub scores correlated modestly with case counts (*ρ*=0.35–0.42), with 82–88% of topological criticality variance independent of epidemic intensity
4. **Non-monotonic bridge evolution:** Transmission bridges did not progressively consolidate but varied non-linearly (2023: 181; 2024: 123; 2025: 192), suggesting complex structural dynamics independent of simple case burden

### 4.2 Mechanistic Interpretation: Stability and Simplification

#### 4.2.1 Network Architecture Resilience

The 1.4-fold increase in edges from 2023 (58,527) to 2024 (81,646) tracked proportionally with municipality increase (4,987 to 5,507), maintaining stable network density. This resilience suggests transmission network formation follows consistent principles across epidemic intensities—municipalities connect based on intrinsic mobility and geographic factors rather than epidemic-driven rewiring.

The stable average degree ( 23–30) across all years indicates that most municipalities maintain consistent numbers of transmission partners regardless of total case burden. This architectural stability contrasts sharply with expected network densification during hyperendemic transmission.

#### 4.2.2 Topological Simplification During Hyperendemicity

Paradoxically, the hyperendemic 2024 year exhibited the simplest topology (274 critical points, 5.0% of nodes) compared to moderate-transmission years (2023: 449 points, 9.0%; 2025: 414 points, 8.2%). This inverse relationship may indicate that high case burdens homogenize the transmission landscape—when disease is ubiquitous, fewer municipalities emerge as structurally distinctive.

The reduction in maxima from 115 (2023) to 56 (2024) suggests fewer localized outbreak sources during hyperendemic transmission. Rather than multiple independent epidemic foci, 2024 transmission appears more spatially uniform, potentially reducing topological gradients that generate critical points.

#### 4.2.3 Bridge Evolution and Variability

Transmission bridges varied non-monotonically: 2023 (181) → 2024 (123) → 2025 (192). This non-linear pattern suggests bridge identification depends on complex spatial-temporal dynamics beyond simple case counts. The 2025 surge in bridges (192, highest across all years) despite similar case volume to 2023 indicates evolving connectivity patterns potentially influenced by:

• Changing mobility patterns or transportation infrastructure
• Serotype replacement affecting spatial transmission dynamics
• Temporal data collection effects (partial year capturing different epidemic phases)

### 4.3 Public Health Implications: Structure-Based Rather Than Volume-Based Targeting

The findings suggest intervention strategies should focus on structural criticality rather than epidemic intensity:

#### 4.3.1 Priority Targeting Principles

**Focus on critical municipalities regardless of case burden:**

• **Structural identification:** Deploy discrete Morse theory annually to identify current-year critical points, as bridge distributions vary non-monotonically (181 → 123 → 192)
• **Intensity-independent targeting:** Maintain surveillance and intervention capacity at critical municipalities regardless of total case counts, since structural importance diverges from epidemic volume (*ρ*=0.35–0.42)
• **Bridge prioritization:** Concentrate resources on identified saddle municipalities, which constitute 40–46% of critical nodes and mediate inter-regional transmission
• **Adaptive allocation:** Adjust intervention distribution based on annual critical point counts (274–449 municipalities) rather than assuming fixed regime patterns

#### 4.3.2 Operational Recommendations Network-informed surveillance

• **Annual topology assessment:** Conduct network analysis each epidemic season to identify evolving critical municipalities
• **Hub-centric monitoring:** Deploy enhanced surveillance (genomic, serological, entomological) at municipalities with hub scores *>*1.9
• **Early warning systems:** Monitor critical points for transmission initiation signals, as their structural position enables early outbreak detection
• **Resource optimization:** During hyperendemic years with simplified topology (e.g., 274 critical points), concentrate resources on fewer high-leverage targets

#### 4.3.3 Adaptive Surveillance Framework

Public health systems should implement annual topological assessment to identify structurally critical municipalities:

1. **Annual network construction:** Use early-season case data to build transmission networks based on documented importation and temporal correlations
2. **Topological analysis:** Apply discrete Morse theory to identify critical points (expected range: 270–450 municipalities)
3. **Structural targeting:** Prioritize resources to municipalities with hub scores *>*1.9 regardless of absolute case burden
4. **Dynamic monitoring:** Reassess critical point distributions seasonally to detect evolving structural patterns

### 4.4 Comparison with Volume-Based Targeting Approaches

Traditional dengue response prioritizes high-incidence municipalities for resource allocation. Our findings demonstrate this volume-based approach systematically misses structurally critical locations. In 2023, the 186 identified transmission bridges had moderate case counts (median range: 229–584 cases, Table 2), placing them outside the top 25% for volume yet exhibiting disproportionate network influence (mean hub score 2.08 vs. 0.47 for non-bridges).

Quantitative comparison suggests topological targeting offers resource efficiency advantages. However, volume-based and topology-based approaches should complement rather than replace each other. High-volume municipalities require intervention for humanitarian and clinical burden reasons regardless of network position. Optimal strategies likely combine volume thresholds (e.g., *>*500 cases/100,000) with topological prioritization among eligible locations.

### 4.5 Methodological Contributions and Generalizability

#### 4.5.1 Validation of Topological Robustness

This study provides the first multi-year validation of discrete Morse theory across epidemic regimes with stable network architecture but varying critical point distributions.

The method successfully identified structurally critical municipalities across networks with consistent density ( 0.0025) but different topological complexity (274–449 critical nodes), demonstrating flexibility in detecting structural heterogeneity independent of connectivity magnitude.

Previous TDA applications in epidemiology focused on single time points or limited temporal ranges. Our three-year comparison establishes that topological criticality identification remains stable and interpretable across fundamentally different epidemic intensities while maintaining consistent network architecture.

#### 4.5.2 Structural vs. Volume-Based Identification

The consistent modest correlation between case counts and hub scores across all regimes (*ρ* = 0.35–0.42) validates a core premise: topological criticality differs from epidemic intensity. Municipalities with moderate incidence (80–584 cases) achieved saddle classification through strategic network positioning rather than volume.

This finding contradicts volume-centric approaches that allocate resources proportionally to case counts. Structural methods identify high-leverage targets that may require fewer resources while achieving greater epidemic disruption.

#### 4.5.3 Transferability to Other Pathogens

The demonstrated regime adaptivity suggests broader applicability:

• **Vector-borne diseases:** Zika, chikungunya, malaria—diseases with similar mobility-driven spread
• **Respiratory pathogens:** Influenza, COVID-19—during periods with varying transmission intensity
• **Sexually transmitted infections:** Network-based transmission with varying contact density
• **Veterinary epidemiology:** Livestock disease spread through trade networks

The key requirement is availability of spatiotemporal case data enabling network construction. The method inherently adapts to whatever connectivity structure exists.

### 4.6 Limitations and Future Directions

Several limitations warrant consideration:

#### 4.6.1 Data Limitations

1. **Reporting completeness:** SINAN depends on passive surveillance; underreporting may vary spatially and temporally. Asymptomatic dengue infections are not captured.
2. **2025 partial data:** Seven-month 2025 data (January-July) may not capture full-year dynamics or seasonal variations. Direct comparisons with full-year 2023 and 2024 data should be interpreted cautiously, though the partial-year analysis still demonstrates methodological applicability across different temporal windows.
3. **Importation data:** Only 22,301 of 1.51M cases (2023) had explicit source documentation; remainder inferred from temporal correlation

#### 4.6.2 Methodological Considerations

1. **Edge construction:** Temporal correlation-based edge inference supplements documented importation data; alternative approaches (genomic validation, mobility data) could refine network structure. Network density remained stable (0.0024– 0.0027) across years, validating consistent threshold application.
2. **Scalar function choice:** Composite risk scores combine multiple factors; alternative formulations might yield different critical point distributions
3. **Normalization artifacts:** Sparse networks may spuriously elevate low-volume municipalities; minimum-case thresholds (25) mitigate but don’t eliminate this
4. **Temporal aggregation:** Annual networks obscure sub-annual dynamics and seasonal variations

#### 4.6.3 Future Research Directions

1. **Genomic integration:** Validate inferred transmission edges using viral phylogenetics and sequencing data
2. **Mobility data:** Incorporate cellular mobility records to refine edge weights and directionality
3. **Sub-annual analysis:** Construct monthly or bi-weekly networks to track topological evolution within epidemic seasons
4. **Intervention studies:** Prospectively test whether bridge-targeted interventions reduce epidemic spread relative to volume-based strategies
5. **Mechanistic modeling:** Integrate topological features into mechanistic transmission models (SEIR, agent-based) to understand causal pathways
6. **Multi-pathogen comparison:** Apply framework to Zika and chikungunya to assess pathogen-specific topology
7. **Climate integration:** Examine how climatic drivers influence network formation and topological structure
8. **Machine learning prediction:** Train supervised models using topological features to enable operational early warning systems

## 5 Conclusions

This study establishes discrete Morse theory as a robust tool for identifying critical transmission infrastructure in dengue networks across varying epidemic intensities. Analysis of three Brazilian epidemic regimes (2023–2025) demonstrates network architecture remains remarkably stable despite 4.3-fold case variation, with network density consistently near 0.0025 (range: 0.0024–0.0027) across all years. Contrary to expectations, hyperendemic transmission simplifies rather than elaborates network topology—the highest-burden 2024 year exhibited the fewest critical points (274) while moderate-transmission years showed greater topological complexity (2023: 449; 2025: 414).

Key validated findings include: (1) topological criticality diverges substantially from epidemic volume, with hub scores showing only modest correlation with case counts (*ρ*=0.35–0.42); (2) critical municipalities show strong statistical significance (Cohen’s d=4.2–6.8) reflecting intentional separation by composite hub score design; (3) transmission bridges vary non-monotonically (181 → 123 → 192) independent of case burden, indicating complex structural evolution; and (4) network density stability (range: 0.0024– 0.0027) despite major case fluctuations suggests transmission pathways follow consistent geographic and mobility patterns rather than epidemic-driven rewiring.

As dengue continues expanding geographically and fluctuating in intensity across endemic regions, topological surveillance provides essential capabilities for understanding transmission dynamics. Public health systems should implement annual network monitoring to identify structurally critical municipalities regardless of epidemic phase, adapting intervention strategies to detected critical point distributions (274–449 targets). Beyond dengue, this methodological framework generalizes to other infectious diseases exhibiting network-driven transmission, offering a flexible analytical tool for network epidemiology.

## Ethics Statement

This study analyzed aggregated, de-identified public health surveillance data from Brazil’s Ministry of Health through the Sistema de Informação de Agravos de Notificação (SINAN). No individual patient identifiers were accessed or analyzed. The study utilized data collected as part of routine national disease surveillance activities mandated by Brazilian public health regulations (Portaria MS/GM n° 204, February 17, 2016). Under Brazilian research ethics regulations (Resolution CNS 466/2012), analysis of anonymized surveillance data for public health purposes does not require institutional review board approval or informed consent. All analyses complied with Brazilian data protection legislation (Lei Geral de Proteção de Dados, LGPD).

## Acknowledgments

We thank Brazil’s Ministry of Health for providing access to multi-year SINAN data and municipal health departments for sustained surveillance collaboration. We acknowledge the value of longitudinal data in enabling robust temporal validation across epidemic regimes.

## Data Availability

Aggregated municipality-level datasets and complete analysis code will be made publicly available upon publication at a permanent repository (available at github.com/trunci/dengue-topology-analysis), as well as network construction protocols, discrete Morse theory implementations, and threshold selection algorithms. Raw SINAN data are available through Brazil’s Ministry of Health data request system (https://opendatasus.saude.gov.br/) following standard procedures.

## List of Abbreviations

BC: Betweenness centrality
DC: Degree centrality
DENV: Dengue virus
DMT: Discrete Morse theory
IBGE: Instituto Brasileiro de Geografia e Estatística
IQR: Interquartile range
OR: Odds ratio
ROC-AUC: Receiver operating characteristic - area under curve
SCC: Strongly connected component
SINAN: Sistema de Informação de Agravos de Notificação
TDA: Topological data analysis
WHO: World Health Organization

